# “Your womb, your choice!” Making an informed decision regarding the timing of pregnancy following miscarriage

**DOI:** 10.1101/2020.09.10.20191858

**Authors:** Faizan Shah, Sohinee Bhattacharya, Kathleen Lamont, Heather May Morgan

**Author notes:** **Corresponding author:** Dr Sohinee Bhattacharya, Dugald Baird Centre for Research on Women’s Health, Aberdeen Maternity Hospital, Aberdeen AB25 2ZL. **Declaration of interest: None**. **Author contributions:** Conceptualisation- SB; Data collation and analysis – FS and HMM (Mumsnet) KL (PIL); first draft - FS; edit and contribute to final draft – all.

## Abstract

The ideal interpregnancy interval (IPI) following a miscarriage is controversial as the World Health Organization (WHO) advise women to delay pregnancy for at least six months. Subsequent research has found that IPI less than six months is beneficial for both mother and baby. The impact of this guidance on the decision-making process for couples/women in this predicament is unknown.

Views of women regarding the optimum IPI following miscarriage were investigated using a thematic framework applied to discussion threads from a popular online forum, Mumsnet (https://www.mumsnet.com). A systematic search of all online information was also undertaken to identify all relevant patient information regarding conceiving another pregnancy after a miscarriage. The findings from the search were tabulated and analysed in relation to the themes identified from the discussion threads on Mumsnet. Ninety-four discussion threads were included. Women saw no reason to wait if they felt ready. Women posted about their frustrations at the multiple sources of conflicting advice they received, at the lack of professional sympathy and felt that being told to wait before trying to conceive after a miscarriage was outdated advice. However, these findings were not corroborated by the patient information currently available online. All web-based patient information gave consistent advice – to wait for at least one normal period before trying to conceive again after a miscarriage and to try for another pregnancy when they felt physically mentally and emotionally ready. None advised waiting for six months. This study highlights that sometimes despite contradictory clinical advice, women are keen to make their own decisions regarding reproductive choice. These decisions are often empowered by peer support and advice which women trust over inconsistent information received from healthcare professionals. In this case, health information appears to have been updated in response to women’s choice rather than the other way around.

**Research highlights:**

- A knowledge gap exists in relation to the views of women regarding the ideal interpregnancy interval following miscarriage.
- This study provides insights into the views and beliefs of women regarding the IPI following miscarriage
- The dominant themes emerging from the study were: there is an array of conflicting advice being provided; there is no requirement to wait following a miscarriage; and the right time is when a couple feels physically, mentally and emotionally ready to try for another pregnancy.
- Current web-based information for patients does not endorse the WHO’s guidance of waiting for at least six months before trying to conceive again after a miscarriage.

## Introduction

Miscarriage, or the spontaneous loss of pregnancy in the first twenty weeks of gestation, affects almost one in five pregnancies (Regan et al., 2019). Loss of a wanted pregnancy is a shocking and sad experience. In an effort to continue family planning, many often inquire about how soon they can try to conceive again. Current World Health Organisation (WHO) guidelines recommend waiting at least six months after an abortion or miscarriage before trying to conceive again in order to prevent adverse outcomes in the next pregnancy (WHO, 2005). This recommendation was based on a single large-scale study conducted in Latin America (Conde-Agudelo, 2004). Although this study was population based and combined data from several Central and South American countries, the history of previous miscarriage was self-reported and therefore could include induced abortions. Subsequently, several other population based primary epidemiological studies (Aref-Adib et al., 2008; Love et al., 2010; DaVanzo et al., 2012) and a systematic review (Kangatharan, Labram and Bhattacharya, 2017) showed that interpregnancy interval (IPI) of less than six months following miscarriage was not associated with adverse outcomes in the next pregnancy and, in fact, may result in better outcomes than delaying pregnancy in direct conflict with the recommendations of the WHO (WHO 2005).

While the academic controversy around the optimum timing of the next pregnancy following miscarriage continues, it is important to understand how the key users of this advice, the women who have suffered a miscarriage, feel regarding this issue. Many couples are faced with the challenge of deciding when to try to conceive again following miscarriage, amidst their immense grief and acceptance processes.

Although about one in five women of reproductive age has experienced at least one miscarriage (Corbet-Owen & Kruger, 2001; Kluger-Bell, 1998), only in the past three decades has the subject been investigated in the disciplines of social studies and psychology (Moulder, 1998). In a systematic review of qualitative research, Dyer et al (2019) explored the decisions regarding pregnancy following perinatal loss. One of the themes identified in this review was directly related to the timing or planning of the subsequent pregnancy. Eleven out of the fifteen studies included in this review reported on this theme. The study participants all felt that the timing of the next pregnancy should be a personal decision based on individual circumstances and experience. Women often spoke about listening to their bodies around deciding when they were ready to conceive again. They also spoke about the inconsistency of advice given by health professionals. While these views are about perinatal loss in general, many of them were also applicable to conceiving after miscarriage or early pregnancy loss.

Qualitative research carried out to date has mainly focused on the period immediately following pregnancy loss or on the outcomes of the subsequent pregnancy. This research study was designed to explore the views and beliefs of couples on the ideal interpregnancy interval following a miscarriage and to systematically search for the current advice given by health care professionals regarding this issue.

## Methods

### Mumsnet Study

#### Design

This study employed a qualitative content analysis (Mayring, 2019) of secondary data (Skea 2008). It utilised a thematic framework applied to online discussion threads on the topic of the ideal interpregnancy interval following miscarriage. The aim of analysis was to generate an understanding of the views and beliefs of individuals and online discussion communities regarding the optimal time to try to conceive after a miscarriage.

#### Setting

In order to gain insights into the views and attitudes of couples towards the interpregnancy interval from a ‘naturalistic setting’, a popular online forum, Mumsnet, was chosen as the primary resource for data collection. Mumsnet is the biggest online forum used by parents in the United Kingdom and around the world; it boasts more than twelve million views per month (Mumsnet, 2018). Mumsnet was launched in 2000 and was advertised as a forum that “Offers product reviews and parenting tips by parents for parents” (Mumsnet, 2018).

The Mumsnet forum allows members to begin a thread or discussion on any chosen topic, as well as to add their comments to existing threads. Furthermore, non-members are also able to search the archived threads on the site using search terms and keywords. The views and beliefs of women were examined using qualitative analysis of specific threads on this popular online forum. The threads were identified during April-May 2019 and a content analysis was applied.

The discussion threads included within the dataset consisted of posts of participants on the website, Mumsnet, who had experienced miscarriage and were attempting to conceive again or contributors giving advice and peer support on this topic, offering their views, opinions and experiences on the ideal interpregnancy interval.

The inclusion criteria were that they were in the English language, posted in the last twenty years and consisted of threads from women/couples on the subject of interpregnancy interval following miscarriage, whether this was in the form of sharing experiences or advice.

#### Data collection

In order to gain an appreciation and understanding of the volume of the data available on Mumsnet, a scoping search for discussion threads was undertaken. The relevant discussion threads were identified using keywords and terms in order to maximise the sensitivity. The keywords that were utilised as search terms in the archives directory on Mumsnet were, “Pregnancy following miscarriage”, “TTC (Trying to conceive) following pregnancy” and “Interpregnancy interval”. Upon searching the discussion threads, it was determined that an abundance of data was available relating to the research question. The selection of Mumsnet as the forum of choice was mainly due to this fact that there were extensive discussion threads surrounding the interpregnancy interval. Only archived threads from Mumsnet were included in the dataset and there was no input in terms of posts or comments from the researcher, this allowed for a minimal level of intrusiveness in the methodology of data collection, restricted to pseudonymised data in the public domain.

This initial search for discussion threads identified over 200 threads. The threads were then pruned according to the inclusion and exclusion criteria. From the initial 200 threads, 94 threads were found to be specifically relevant to the research question and thus finalised as the dataset for this research.

The responses and discussions in the threads were downloaded from Mumsnet using NCapture (NCapture, 2020) imported, organised and then coded using a thematic analysis framework on the NVivo software into themes and subthemes.

#### Data analysis

The community level beliefs of women on the topic were established by the development of a thematic framework. A sample of six threads from the entire dataset were randomly chosen and double coded. In order to increase reliability, two independent researchers (FS and HM), from differing educational backgrounds undertook the initial coding process for this sub-sample. Throughout each step of the study, discussion was maintained among all authors, in order to minimise potential biases (Primeau, 2003). Thereafter, themes identified in the sub-sample of threads were discussed with the principal investigator (SB) and the other authors, and compared with the literature, and a thematic analysis framework was created for application to the remaining threads in NVivo12 software. The initial separate analysis of the chosen discussion threads revealed that thematic variety as well as similarity existed between the threads. Following the identification of the themes from the threads, key themes were further refined into themes and subthemes and subsequently coded using the NVivo software. Exemplar quotes for each theme were reported to illustrate the type of responses that the threads posts were receiving and also to demonstrate the nature of the theme as it appeared in the actual discussion threads.

#### Systematic analysis of Patient Information Leaflets/ Websites

A systematic search of all Patient Information Leaflets accessible online was carried out using Google search engine with key words such as “miscarriage,” “patient information,” “Early Pregnancy Unit,” “miscarriage support and advice”, “wait time to pregnancy following miscarriage” and “conceiving after miscarriage.” Across the UK, Early Pregnancy Assessment Units (EPAUs) provide care and support for women who suffer miscarriage in early pregnancy. Therefore, in addition, the websites of individual EPAUs in the UK were also searched for any relevant patient information that had not been picked up by the key word search. The findings from the systematic review of PILs/websites were collated, tabulated and analysed in relation to the themes obtained from the discussion threads on Mumsnet.

## Results

The dataset for the Mumsnet based qualitative analysis comprised 94 threads, identified through searches of archived discussion threads on Mumsnet. These 94 threads met the inclusion criteria as they discussed the views and beliefs of women regarding the ideal IPI following a miscarriage. These mainly comprised of the themes: ‘feeling ready’ to conceive again, the widespread conflicting advice women were receiving from healthcare professionals, as well as the main recurring theme where women believed there is no requirement to wait following miscarriage and that one should be able to try for another pregnancy when they felt physically and emotionally ready. Table 1 presents the main themes and subthemes that were initially identified independently and refined into a framework using a sample of 6 threads from the dataset of threads. Following agreement between the independent assessors of the themes the analysis framework developed was applied to the full dataset to allow coding.

**Table 1:** Table of identified themes and subthemes

| Themes | Subthemes |
| --- | --- |
| Reliability of sources of information | 1. Doctors<br>a. Ease for the doctor (in terms of calculating cycles)<br>2. Nurses<br>3. Midwives<br>4. Science<br>5. Online forums<br>a. Mumsnet (positive)<br>b. Google (negative) |
| Conflicting advice |  |
| Traditional practices/norms/old advice | 1. Wait for one period cycle<br>2. Being told to wait by healthcare professionals |
| No need to wait |  |
| Lack of professional sympathy |  |
| Fears form personal/family experience | 1. Taking from too many sources<br>2. Trying too early may lead to another miscarriage |
| Husband/partner/family | 1. Stress caused due to family/friends successfully conceiving<br>2. Relationships |
| Quoting statistics | 1. Desire for a personalised prediction<br>2. Risk – Probability |
| Feeling ready | 1. Physical<br>a. Ovulation<br>i. Feeling<br>ii. Temperature<br>iii. Kits<br>2. Mental<br>a. Try not to focus on conceiving<br>b. Do whatever feels right<br>3. Emotional<br>a. Grief<br>b. Getting over the loss<br>c. Desire to try again |
| Increased fertility following a miscarriage |  |
| Calculating cycles | 1. What is a period vs. miscarriage<br>2. New or old pregnancy |
| Taking supplements/activities | Folic acid (positive) |
| Investigations/scans/ symptoms of pregnancy | 1. Apps for anxiety<br>Scans used as milestones |
| Feeling ready | 4. Physical<br>a. Ovulation<br>i. Feeling<br>ii. Temperature<br>iii. Kits<br>5. Mental<br>a. Try not to focus on conceiving<br>b. Do whatever feels right<br>6. Emotional<br>a. Grief<br>b. Getting over the loss<br>c. Desire to try again |
| Increased fertility following a miscarriage |  |
| Calculating cycles | 3. What is a period vs. miscarriage<br>4. New or old pregnancy |
| Taking supplements/activities | Folic acid (positive) |
| Investigations/scans/ symptoms of pregnancy | 2. Apps for anxiety<br>3. Scans used as milestones |

### Themes (in bold)

#### Reliable Sources of information

Many of the posts analysed were related to the advice received about conceiving after miscarriage. The sources of information were healthcare professionals including doctors nurses and midwives; online information using Google or fora such as Mumsnet.

#### “Doctors’ ease of calculating cycles”

In 35 of the 94 threads that were analysed, conversation regarding being advised to wait by a doctor were attributed to have been based upon an **ease** for the physician to calculate menstrual cycle, rather than being based upon evidence of the harms and benefits of conceiving again following miscarriage. The user ‘frostymorning’ highlighted the fact that they believe doctors giving ‘old advice’ was simply in order to ‘help with dating’ the pregnancy.

#### frostymorning Mon 12-Nov-07 11:07:32 Add message | Report | Message poster

Sorry to hear your bad news, it’s very distressing to think everything’s alright, then it’s not, then it is and then finally it’s all gone wrong. **There’s no reason to wait to TTC (trying to conceive), the old advice was to wait until a period to help with dating but scans are so good now they can date accurately from th**at. Lesley Regan’s book is very good re chances of repeat miscarriage (usually small) and I found it quite encouraging to read her statistics regarding subsequent pregnancies.

Furthermore, another member displayed their frustration at the advice given by doctors and the reason being to allow them to ‘date properly’.

#### The “date properly” thing is ridiculous though.

I charted when I conceived DS (now 4) – knew pretty exactly when I conceived, but the dating scan put conception a week earlier, and that was the date that had to be used. DS arrived a day before I had calculated he was due. **So why wait for a period when it’s only the dating scan that is trusted? The only advantage I can see as regards dating is that it saves you from going for an early scan before 7 weeks** (the magical date for reliably seeing the heartbeat and therefore helping to put your mind at rest).

#### Positive support from online forums

Numerous threads included members expressing their appreciation of online forums in their journey to conceive following a miscarriage. Mumsnet was repeatedly mentioned as a “**positive source of information”** and as a platform for women to support one another and share experiences. It was noted that despite the many professional and online sources of information that are available to mothers, they seemed to prefer discussion threads where they could take from other people’s experiences more beneficial.

#### orange47 Sat 18-Feb-17 20:48:01 Add message | Report | Message poster

May I join? I can’t tell you how much lurking on here has helped over the past few weeks while I have been coming to terms with my MC (miscarriage) and wanting to try again but not rush things too quickly. **You are all so lovely and supportive! It has been so helpful knowing that I am not alone in this, although of course I am sorry any of us has to be here**. We are TTC (trying to conceive) #1, got PG (pregnant) on cycle 3 or 4 (not sure if I count the first one!) and had a MC (miscarriage) at 6+4 early Jan. First AF since has now arrived and it was such a relief, hoping my cycles now go back to normal and we can try again properly this month!

#### Feeling ready

One of the most dominant themes throughout the discussion threads was that, despite the debates on whether one should wait or not before trying to conceive again, many people recommended to try and conceive again when one feels physically, mentally and emotionally ready to do so.

#### skibump Mon 12-Nov-07 22:26:11 Add message | Report | Message poster

There is no evidence that waiting any length of time or not waiting before TTC again makes any difference at all. Current thinking is that you should go for it again WHEN YOU FEEL READY. That may be immediately after a MC, after a period of time or never again, IYKWIM. My understanding is that the advice to at least let 1 period come and go was for dating purposes mainly. Now that we have v accurate scans to date a pregnancy knowing your LMP is not that

##### Hopefulinbalham Fri 13-Oct-17 21:02:06 Add message | Report | Message poster

**I was told very recently that you can try straight away if you feel emotionally ready**. The advice of waiting one month was just for dating purposes but they can estimate the date from the scans anyway. As long as the scans show that everything internally is as it should be, you can go ahead

On the other hand, others took a different standpoint, stating that conceiving straight again following a miscarriage is extremely difficult emotionally and mentally and many of the discussion threads incorporated members emphasising the importance of maintaining one’s mental health, as suffering a miscarriage bears a considerable mental/psychological burden.

#### TinyTear Tue 25-Mar-14 10:45:47

Some people jump straight to it, others need some time to process things. Unfortunately I have had 5 mcs and most time I have taken at least 2 months before trying again& more after number 3 as I was waiting for test results don’t jumpt before you feel ready… **mental health is also very important, not just your body**

Although the advice of waiting before trying to conceive again is small in comparison to the many people advising that there is no need to wait, the reasoning/justification behind this advice is understandable.

#### TinyTear Tue 25-Mar-14 10:45:47

**Boogs I have mixed feelings about ttc again straight away**. After my first loss we didn’t wait at all and I always blamed that as part of the reason we mc again. Having said that though I recently had a mc at 12 weeks. I was determined to wait one cycle but fate intervened and somehow I concieved only 2 weeks later. I am now 23 weeks pg. **So I don’t think there is any real physical reason why you should wait especially if it might take you some time to concieve again. Emotionally it is a different story. You may feel you are ready to try again and just want to move on but it hits you really hard once you have that blue line**. Pg is just never the same after a loss and you become so paranoid and anxious that the first 3 months are really hard going. There is no reason to assume it will happen again but the fear is always there and its this side of pg that you have to be ready to deal with. **The right time is when you feel ready to cope**.

Furthermore, in multiple threads throughout the analysis, mothers stipulated that it is very important to allow the body to heal physically before trying to conceive again.

#### chickenwings Mon 22-Mar-10 19:52:58 Add message | Report | Message poster

Firstly, I am so sorry to hear that you are miscarrying. It is so upsetting, so do give yourself time to grieve if you need to**. I think people tell you to wait to give yourself time to get over it. It is physically tiring just because your hormones are all over the place, however I don’t think there is any reason why your body wouldn’t “be ready” (**though I am no doctor). You won’t get pregnant if your body is not “ready”. Just so you know, I have had 3 miscarriages. The second I had at about 8 weeks and I got pregnant a month later, had my third miscarriage again at about 9 weeks, then fell pregnant again straight away and now have DS3.

#### Conflicting advice

Conflicting advice that women received was identified as a recurring theme throughout the threads. The multiple sources of information, as well as the absence of a well informed and highly regarded guideline on the matter of the ideal interpregnancy interval following miscarriage, there has predictably been a multitude of conflicting advice being provided to women by differing healthcare professionals, online sources as well as experiences of friends and family. Conflicting advice in terms of how long to wait and the harms and benefits of this have been the cause of a great deal of distress and confusion for those trying to conceive following a miscarriage.

#### Sarah295 Tue 25-Feb-14 22:31:31

**There are so many conflicting stories about when you can try to conceive again. The miscarriage association even point out that most doctors say to wait until after your first period and that there is even evidence to suggest that you are less likely to miscarry again if you conceive within the first 6 months**. I do not see how I can begin to get over our loss until I am pregnant again. Two weeks ago I could tell that I was ovulating and pregnancy test was negative – my body wa getting back on track. 2 weeks later I have started my period- it’s heavy but nothing much worse than normal. **What I want to know is whether or not I should even be thinking about trying again this month- I could not cope if it were to happen again but nor could I cope waiting to try and if we struggle to fall pregnant again I will drive myself crazy with**

A common issue related to the lack of a robust evidence base to support the advice given.

#### skibump Mon 12-Nov-07 22:26:11 Add message | Report | Message poster

Hi Twinklewishes. I m/c naturally in May at 7 wks (so not sure if it’s relevant…but) the **doctor initially said I should wait for my next period before TTC (trying to conceive), but when I asked why she really couldn’t answer – and said it was probably just old-fashioned advice to get a fix on dates**. I think you have to give yourself time to get over what’s happened, but you should do what feels right for you(r family) Sorry for what you’ve been thro, it must have been awful. Thinking of you

Receiving conflicting advice from healthcare professionals, whom the public hold in high regard, has been shown to leave women confused and has the affect of causing one to question the integrity of the medical profession. Smartiejake, amongst many others, displayed the manner in which two healthcare professionals were giving her conflicting advice and ultimately it is up to the individual to decide which advice to take.

#### smartiejake Fri 04-Jan-08 10:05:58 Add message | Report | Message poster

I had a miscarriage at 7 weeks. Had to have and D & C as it didn’t happen naturally (they saw it on a scan after I had a small bleed) **We had different advice about when to try again. Nurses said wait 3 months and Dotor said just to wait for next period. I took the doctor’s advice and got pregnant again very quickly**. (The next month would you believe.) Pregnancy went smoothly and dd2 was born about 2 months after when miscarried baby would have been due.

#### Being told to wait is a traditional practice/norms/old advice

A vast majority of mums posting in the discussion threads seemed to have a clear view that the advice to wait before trying to conceive again was largely outdated and old fashioned. The theme of receiving advice from healthcare professionals that was deemed to be old fashioned or in accordance with traditional practices was seen to appear throughout the majority of the threads which were analysed. The old fashioned advice to wait before trying to conceive again is mostly down to dating reasons, however with the advent of scanning and progress in technology, this is said to be no longer required.

#### frostymorning Mon 12-Nov-07 11:07:32 Add message | Report | Message poster

**There’s no reason to wait to TTC (trying to conceive), the old advice was to wait until a period to help with dating but scans are so good now they can date accurately from that**. Lesley Regan’s book is very good re chances of repeat miscarriage (usually small) and I found it quite encouraging to read her statistics regarding subsequent pregnancies. I’m 5 weeks pregnant following a MC (miscarriage) last spring and am a wreck because I’m cosntantly worried it’s all going wrong and I don’t know it. I suppose it’s to be expected really. I can’t really say anything to make you feel okay but I just wanted to respond to your post.

In another thread, responses mirrored these thoughts and women expressed their frustration at the fact that they felt there was inconsistency and a lack of honesty on the part of the healthcare professionals. There was a sense of requiring an official and honest answer to how long to wait following a miscarriage to get pregnant again and not simply advice, which is old fashioned and based on traditional norms.

Hi Ghosty – I’m sorry you are so frustrated. I must be really hard. But fingers crossed for Thursday eh! **I think that women are advised to wait a few months because it is the convention to do so, although I have to admit it does give you time to heal emotionally**. After a loss you are so desperate to move forwards that you can’t really imagine how hard a subsequent pg will be so you do need time to recover a bit emotionally. **I just wish they would be honest about that as it would make it easier to wait. Dating the pg isn’t really an issue with all the scans they do today anyway**

#### Wait for one period cycle

Another dominant theme, which was closely related to the idea of traditional advice, was the idea of waiting for one period cycle before trying to conceive again following a miscarriage. This theme was amongst the top three recurring themes throughout the threads, with multiple women reporting that they had been advised to wait for one period cycle following a miscarriage Although, there were women who deemed this advice as being helpful in terms of the period in between allowing them to be physically, mentally and emotionally ready to try again for a baby. This theme is therefore seen as a continuation of the previous one as being based on traditional advice and practices.

#### Out2pasture Fri 03-Jun-16 06:20:03 Add message | Report | Message poster

being the op lost the baby in her second trimester **physicians recommend waiting 3–4 cycles**.

Furthermore, it was evident through analysis that women had been told to wait for a number of period cycles before trying to conceive again, and further advised that not following this advice could result in a recurrence of this miscarriage for them, however this has not been supported by guidelines or clinical evidence.

#### fnisashah88 Wed 18-Oct-17 05:06:21 Add message | Report | Message poster

Hi. Very sorry about your loss. I had a miscarriage just over 3 weeks ago. **I was told to wait for 2 regular cycles to try again. Also told if I was to get pregnant before that there is a another chance of miscarriage**.

Others were quite cynical regarding the advice to wait before trying to conceive again and went on to have successful pregnancies having ignored the advice to wait a period cycle before trying again.

#### Lozmatoz Wed 18-Oct-17 17:48:27 Add message | Report | Message poster

Yes. Realised had a missed miscarriage at 12 week scan. Had d&c at 14 weeks and **was told to wait to have s period before trying again (because it was easier to date). I ignored that advice and was pregnant again without having a period. He’s 5 now!**

However, on the other hand, a minority of responses advocated the advice of waiting for a period cycle before trying to conceive again. Most of the proponents of this cited dating reasons for advising this, as well as women claiming the time in between is required to heal emotionally, physically and mentally.

#### HappyToBeAlone Mon 13-Jun-11 19:10:46 Add message | Report | Message poster

Jasmine thank you, I just hope i get to have an early scan to set my mind at rest, Im not even sure on my dates either now**, I can see why people wait a cycle as it makes it easier to date I have had 2 MC (miscarriage)’s one in 2006 and the last one last month so i am worried I am going to try and not get excited till i see a scan and a healthy heartbeat pumping away**

#### No need to wait

Given that there were a variety of opinions from doctors, nurses, midwives, family and friends as well as online resources, many women had concluded that there was no medical reason to wait before trying to conceive again. Almost all of the threads had at least one member responding with the advice that there is no need to wait and the fact that one may try again as soon as they feel ready to do so.

#### Squiff70 Tue 05-Mar-19 18:23:06 Add message | Report | Message poster

**You really need to wait for a normal period after a miscarriage because the womb lining needs time to thicken and recover**. I’m sorry for your recent loss and wish you every luck in having a healthy pregnancy in the near future

##### Izzybel Mon 21-Jan-08 20:10:53

sorry to hear about your miscarriage. I **have heard about people conceiving straight after a Add message | Report | Message poster MC (miscarriage) without having a period after**. Has the bleeding stopped, as they don’t recommend that you have sex until it has, because of risk of infection etc? **Also, doctors usually recommend that you wait until you have had one period before trying to conceive, I think this is to make it easier to work out dates etc. I had a MC (miscarriage) in July 2006 and got pregnant again in the September, so I think you can be pretty fertile after. As long as you feel ready to start trying again I wouldn’t have thought there was any reason not to. If your body is ready to conceive it will**

It was observed that, upon doing research, women found that there was no medical reason to wait before trying to conceive again. This is an example of someone who has done their research and has concluded the main points of advice, separating what is recommended from what is supported by evidence.

#### cornflakegirl Mon 27-Apr-09 13:11:16

I have had three consecutive miscarriages. With the first and the third, I conceived again without waiting for a proper period. I’m currently 5 months pregnant. **Personally, I saw no reason to wait to conceive – the research I did suggested no medical reason**.

#### Lack of professional sympathy

A lack of sympathy on the part of professionals dealing with women who have suffered a miscarriage was an issue which caused a great deal of distress. From condescending remarks to unempathetic judgements, women in such situations have been made to endure added suffering. Uncompassionate professionals have caused women to question the integrity of the profession and have resulted in women feeling uncomfortable in confiding in their healthcare professional. This was also displayed in the research conducted by Anderrson et al, who found that women felt a sense of vulnerability and lack of professional support following miscarriage. (Andersson, Nilsson and Adolfsson, 2011)

#### Flatwhite31 Sun 22-Oct-17 21:39:46 Add message | Report | Message poster

I have absolutely no symptoms before my periods, and they have only lasted 2 days**As for the midwife, my midwife was so lacking in compassion I wrote an official complaint (as advised by my GP, who was extremely shocked at the poor care) so there is absolutely no way I’m going to confide anything in them. They were utterly useless, and I’ve lost all trust in them**.

As well as a lack of sympathy, some women have experienced an unforgiving lack of support and guidance following a miscarriage. Struggling to gain support from healthcare professionals, mums have resorted to online threads such as Mumsnet and have displayed their appreciation of the support and guidance that other women in similar scenarios have given them.

#### user1480930113 Tue 14-Feb-17 08:08:51 Add message | Report | Message poster

Thanks all, you’re all a lovely lot! She does know about my MC (miscarriage) I shouldn’t be so eager and as you say let my body do its thing and recover. **The hospital didn’t actually tell me anything after I have gathered my info off here x**

#### Fears from personal/family experience

Miscarrying a pregnancy is a highly emotional experience, which has shown to cause a great deal of anxiety and fear. Women who have had previously suffered a miscarriage are increasingly apprehensive in future instances due to a natural but sometimes unwarranted anticipation that the miscarriage may recur. As well as fears due to their own personal experiences, women are haunted by stories of others who did not go on to have successful pregnancies following a miscarriage which again adds to their anxiety and scepticism.

Hi tashm I am sorry for your loss. I lost my first baby at 17 weeks last year, and both my GP and my gynaecologist said I could ttc as soon as the bleeding had stopped. **I believe in most cases there is no reason to wait for 3 months. We tried straight away and conceived again after 3 months**. Best of luck xx

##### GandTnow Tue 25-Mar-14 16:02:00

So sorry to hear of your loss. Add message | Report | Message poster I had a MMC (missed miscarriage) at 8 weeks, found out at the 12 week scan. Had an ERPC and then some issues with cycles. **All this started in November and it has taken me a long time to feel back to ‘normal’. I still feel very sad sometimes and find it difficult seeing women who are pregnant, I’m very aware that the baby I lost would be due in May and I ache to have that baby now and be planning the room and looking at toys etc. That being said, I didn’t feel ready to try again straight away. I needed time to sort myself out first**.

#### Trying too early may lead to another miscarriage

The notion that trying to conceive too early following a miscarriage increases the risk of a recurrent miscarriage was evident in the threads and was one of the main reasons why women contemplated waiting for a period of time before trying to conceive again. As for women who fell pregnant soon after a miscarriage, they are challenged with the constant anxiety and guilt that trying too early may have sabotaged their chance at a successful pregnancy once again.

#### maxine412 Mon 16-Oct-17 10:08:29 Add message | Report | Message poster

**So would you say I’m more likely to lose this baby because I’ve fallen pregnant straight away?**

##### Jaraa Thu 04-Jan-18 22:27:46 Add message | Report | Message poster

Hi everyone, I m very worried.. **My experience in the hospital was horrible. It was hard physically, mentally and I begin to feel better. For a few days, I feel pregnant. I will be happy but not ready, I am still afraid and anxious after**

However, a number of members participating in the discussion threads were quick to clarify this issue with statistics and facts.

#### CorrieDale Wed 23-Aug-06 07:13:36 Add message | Report | Message poster

**Stop worrying! You are no more likely to have a m/c now simply because you’ve already had one. In fact, even when you’ve had three, you’re still more likely to have a child than another. Statistically speaking. Smoking and caffeine are more likely to ‘cause’ a m/c than a previous m/c**. I know you’ll worry anyway, because you can never have another stress-free pregnancy once you’ve m/cd. But try not to. **Read up on the subject – there’s loads of stuff on the net - I found the statistics quite reassuring**.

#### Stress from husband/partner

As well as its effect on the mother, loss of a pregnancy has a lasting emotional and mental effect on the father of the baby. Most of Mumsnet’s women portrayed that their partners have been extremely supportive and caring throughout their ordeal. The presence of a partner throughout the process has helped women mentally and emotionally as well as allowing them to heal and recuperate comfortably.

#### PineapplePol Wed 13-Jun-12 11:29:04 Add message | Report | Message poster Like

**I’ve found confiding in a close friend useful as having extra emotional support is really helpful**. Just being able to express your fears and concerns helps - and its not always easy to do with your DP (darling partner) following MC (miscarriage) as they have their own concerns. I know mine battened down the hatches emotionally to begin with. **Now we do talk to each other most nights about how we are feeling and the mutual support really helps**.

However, numerous women also expressed the stress and pressure that are caused by their partners, as well as a lack of empathy and understanding. During this delicate period of a woman’s life, support from the partner is of utmost importance and being sensitive to one’s feelings and emotions is paramount.

#### Twinklewishes Tue 13-Nov-07 09:49:03 Add message | Report | Message poster

Thankyou everyone for your messages of support. I feel like the balloon left after a party that is waiting to pop. It is all starting to happen now. **My husband has started making terrible comments about it all and I can not cope. According to him, I did not need any flowers as my mother had sent some already**. And he amazed everyone when he thought that buying be a bag of donuts (as they were the cheapest thing in the supermarket(his words)) were suficient. Eventhough he was told I would be on a drip for three days and could not eat. **It just could not get any worse. And since then he has told me that he has paid out enough as he works and I don’t (can’t, as I look after my other children). I am just feeling very low and wanted to get things off my chest. I always thought that he was a caring person eventhough he always gets it wrong, but this time he said some things that he can not take back this time**.

#### Stress caused by family/friends successfully conceiving

Successful pregnancies occurring around someone who has suffered one/multiple miscarriages is difficult for one to experience. Women expressed throughout the threads that successful pregnancies of family and friends would serve as a bitter reminder of what they could not do successfully. Some participants reported feelings of obsessiveness and envy towards other, being built up due to the miscarriage they have suffered.

#### Sunseeker17 Mon 13-Feb-17 08:51:54 Hello everyone Add message | Report | Message poster

I’m starting to feel like I might not ovulate this cycle (I suppose my body will know when it’s ready but just quite frustrating). **I’m trying not to obsess about it but it’s very hard. I feel like suddenly EVERYONE around me is pregnant. The pregnancy related things I used to think about in my down time at work and at home- all that excitement- it’s all gone and it’s left a big hole in my life. I know it sounds incredibly irrational but in my darkest moments I’m starting to feel like it will just never my destiny to be a mother**.

#### Quoting statistics

Due to the mass disparity and confusion in opinions and the conflicting advice on when to try and conceive again following a miscarriage, a number of people preferred posting statistics to estimate their chances of success. Using statistics and risk ratios to explain chances to a mother has been shown to have a reassuring effect. Furthermore, there is seen to be a requirement for a personalized prediction of success, which is not currently available.

#### Midgetm Wed 13-Jun-12 08:53:34 Add message | Report | Message poster

Pregnancy after miscarriage is harder, because the innocence is gone, but you just have to take each day at a time. **Take comfort in the statistics that one miscarriage does NOT mean that you are more likely to have another**.

#### Increased fertility following a miscarriage

Many proponents of trying to conceive again as soon as possible have stated that the body is in the ideal state to conceive again immediately following a miscarriage i.e. fertility is highest at this point. There has been a magnitude of research conducted on the hypothesis that women are most fertile immediately following miscarriage, which has yielded conflicting results. There is much doubt evident even in amongst the Mumsnet community concerning increased fertility following a miscarriage. Healthcare professionals have been stated to have declared this as a myth.

#### ouzanne Mon 05-Feb-18 11:27:21 Hi all, Add message | Report | Message poster

**I asked my gyne about increased fertility after miscarriage and he said that this was a complete myth/old wives tale**.

#### Calculating cycles

##### New or old pregnancy

An issue which was a seen as a cause of a great deal of confusion and uncertainty is the inability to distinguish between an old and a new pregnancy. This may be due to the fact that hormonal levels may stay elevated and symptoms may remain even after a miscarriage therefore not allowing women to plan a new pregnancy or be certain on whether they are pregnant again or not.

#### Char1230 Tue 19-Jun-18 08:33:36 Add message | Report | Message poster

**Is there any chance me having my MC (miscarriage) during my period and falling pregnant again on the second ovulation. Im very confused**.

##### Taking supplements

A large number of the threads included in the dataset had members advocating the use of supplements and undertaking of certain activities in order to increase the chances of success in pregnancy following a miscarriage. One of the most popular and frequently recommended supplements was folic acid, which is also scientifically proven to be advantageous in pregnancy.

#### emily86 Mon 13-Feb-17 10:15:30 Add message | Report | Message poster

**I just take bog standard folic acid. Tried pregnacare for a while but found they messed with my cycle and I’d spot for 5-7 days before my period started. I too have always had a break from folic acid whenever I’ve had a miscarriage**. After this last one I’ve started taking **Ubiquinol as it can help improve egg quality but it is quite pricey**. And take **vit D** when I manage to remember

##### Investigations/scans/symptoms of pregnancy

Rather than relying on arbitrary arguments and conflicting opinions regarding how long to wait following a miscarriage to try and conceive again, women have turned to more practical and evidence based ways of measuring their readiness/pregnancy/symptoms of pregnancy to plan their pregnancies. With the advent of state of the art scanning techniques and investigations, there is no longer any need to make a decision based on opinions, which lack an evidence base.

As well as being utilised for dating pregnancies and determining viability, investigations are used as reassurances for women during the process of trying to conceive again as well as in a new pregnancy where they are used as milestones to ensure the pregnancy is progressing in a healthy manner according to its corresponding dates.

#### WelshMammy123 Sat 06-Jan-18 17:29:30 Add message | Report | Message poster.

I know you asked about an early **scan up thread - maybe that’s worth Petunia our local hospital offers a scan at 8wks if you have had a previous m/c and then the regular ones at 12 and 23wks. I was wondering whether or not to accept the 8wk scan as my m/c was after this anyway, but I think as most of you are doing I will accept it as it an be like anther milestone if the
baby makes it that far and then the next milestone is 12wks, 16wks and so on**. Taking each day as it comes. Although I’m sat here today wondering if its viable now at 5wks.

I had a miscarriage in May and another in September, now 17 weeks pregnant. **It’s incredibly tough and I haven’t mastered the anxiety yet, despite many reassurance scans!** it helped me to know the key times when chance of miscarriage reduces (stats are on the miscarriage association web site) and to have **private scans at those points - think I’d have gone mad without them!**

##### Feelings of guilt

Following a miscarriage, women occasionally experience guilt and tend to blame themselves or their actions for the loss of the pregnancy. This has the effect of not allowing optimism and confidence in women to try again as they are still in grievance/mourning their previous loss.

#### boogs Mon 14-Apr-03 17:18:00 Add message | Report | Message poster

**Having said that I did and do still feel a great sense of loss and cant help feeling that it was something I did that caused the m/c**.

##### Theme frequency

The themes developed in the thematic framework were used to organise and interpret the contents of the posts. The most dominant theme throughout the thematic analysis of the threads was the advice that women should try and conceive again once they feel “Physically, mentally and emotionally ready” to do so. Many women believed and repeatedly stated that the advice to wait was based on ‘Traditional norms and old practices’. The frequency of the various themes is illustrated in the hierarchy chart presented below. (See figure 1)

**Figure 1:**
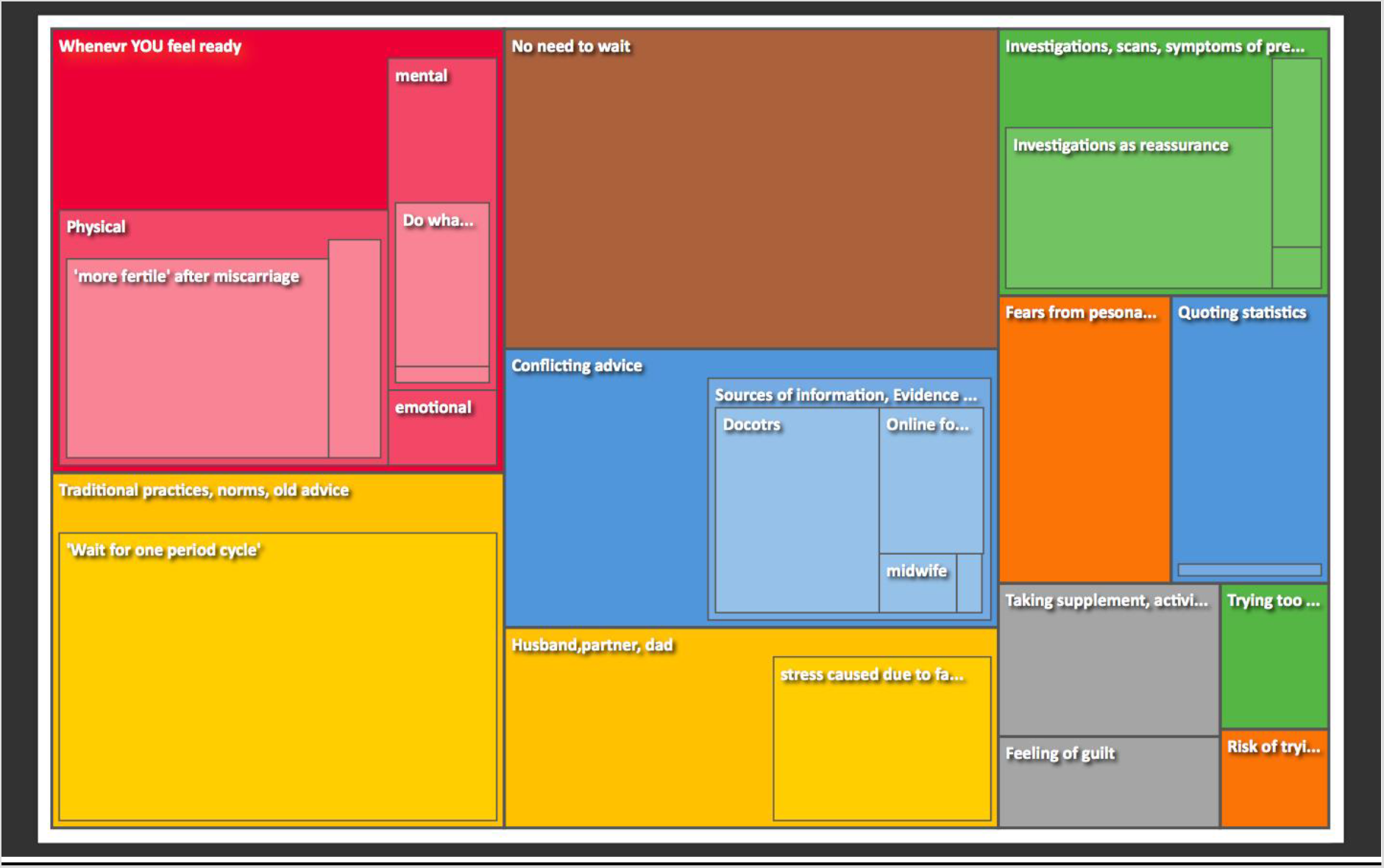
Hierarchy chart illustrating frequency of themes appearing throughout the threads

##### Advice given to couples in Patient Information Leaflets

The results of the search for patient information regarding timing of pregnancy following miscarriage are presented in Table 2. Forty eight PILs were identified; six from national organisations (professional bodies or national charities), 35 from individual hospitals or health boards in the UK and seven from national organisations and hospitals in other countries. The information leaflets were dated between 2011 and 2020 (see Table 2). Nine leaflets (all but one of these were from hospitals in the UK) did not provide information regarding timing of pregnancy following a miscarriage, 17 advised that there is no need to wait, 20 advised women to wait for at least one normal period before trying to conceive again so that accurate dating of the subsequent pregnancy was possible, and two (both from hospitals in the UK) advised women to wait for two to three normal periods before trying to conceive. Leaflets also warned that fertility returned almost immediately after a miscarriage and advised against having sexual intercourse before the miscarriage bleeding had stopped in order to prevent infection. Many of the information leaflets or brochures left it up to the couples to make the decision themselves, saying “*Once the miscarriage is over, you can try to get pregnant again as soon as you and your partner feel ready*”. None of the leaflets advised couples to wait for at least six months as per WHO guidance (WHO 2005). Some of the leaflets (Miscarriage Association UK, NICE Clinical knowledge Summary and Miscarriage support New Zealand) directly cited published academic papers (Love et al., 2010 and Kangatharan et al 2017) as the evidence base for their recommendation but most of the others did not include any citation or bibliography.

**Table 2.**
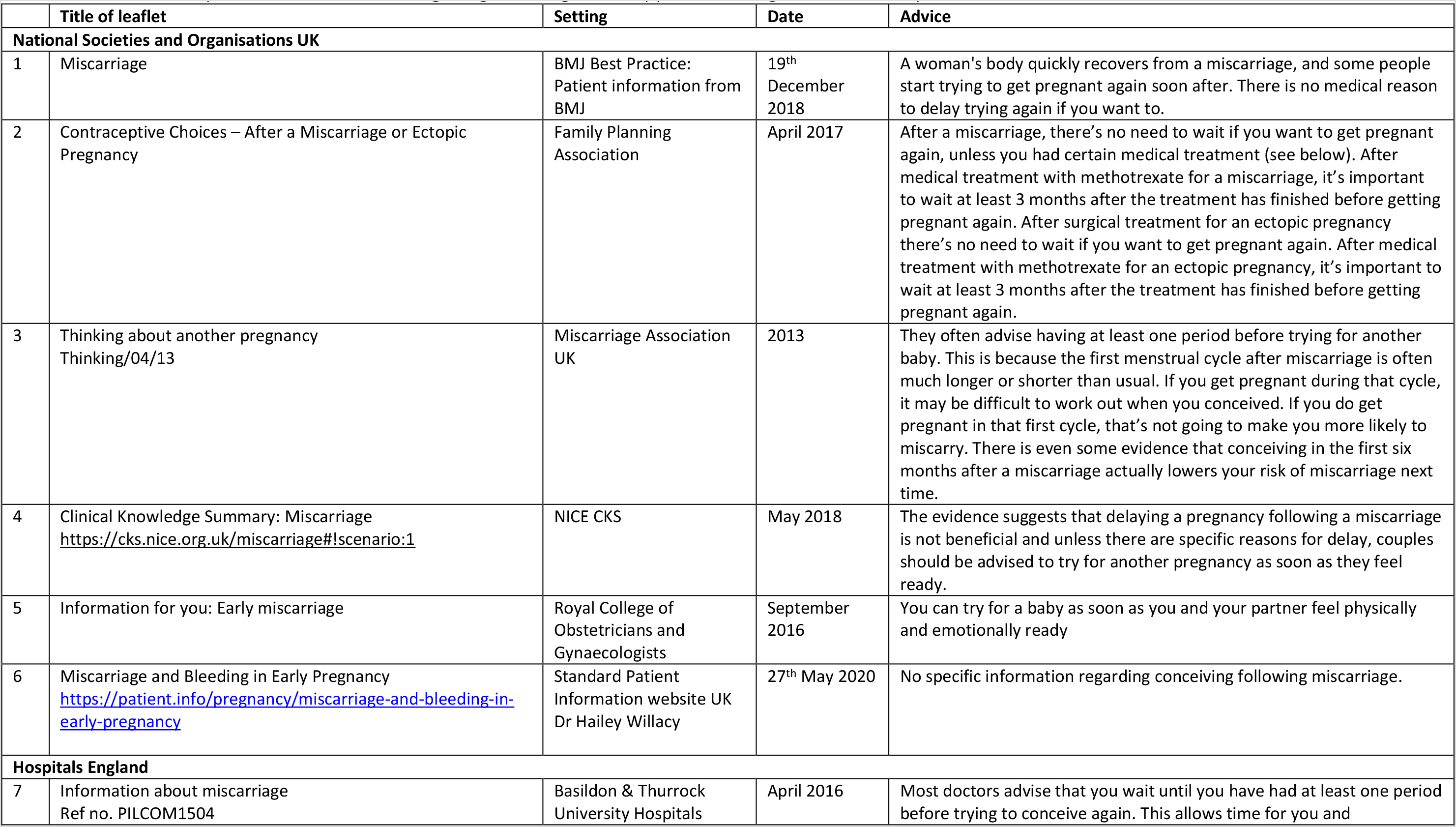
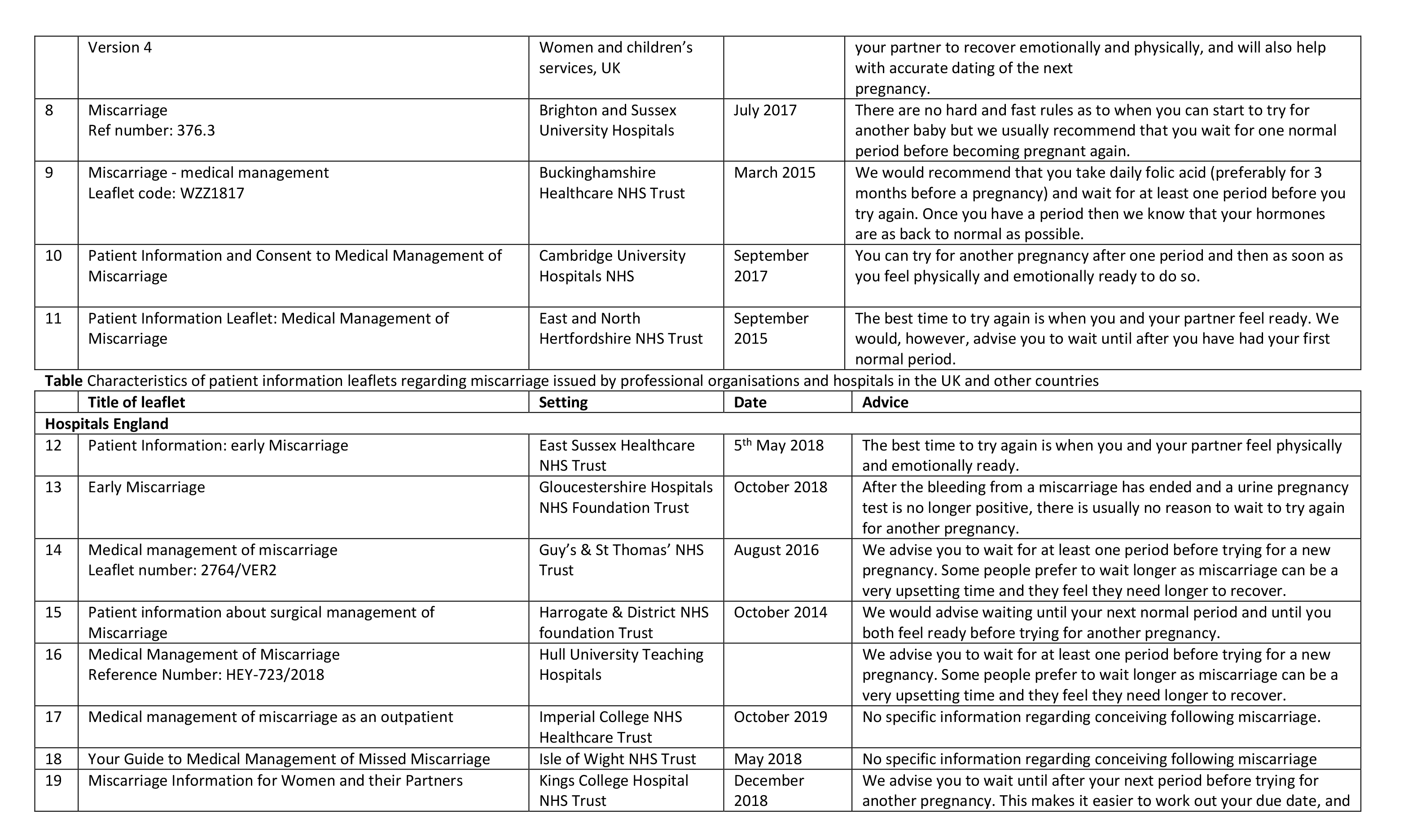
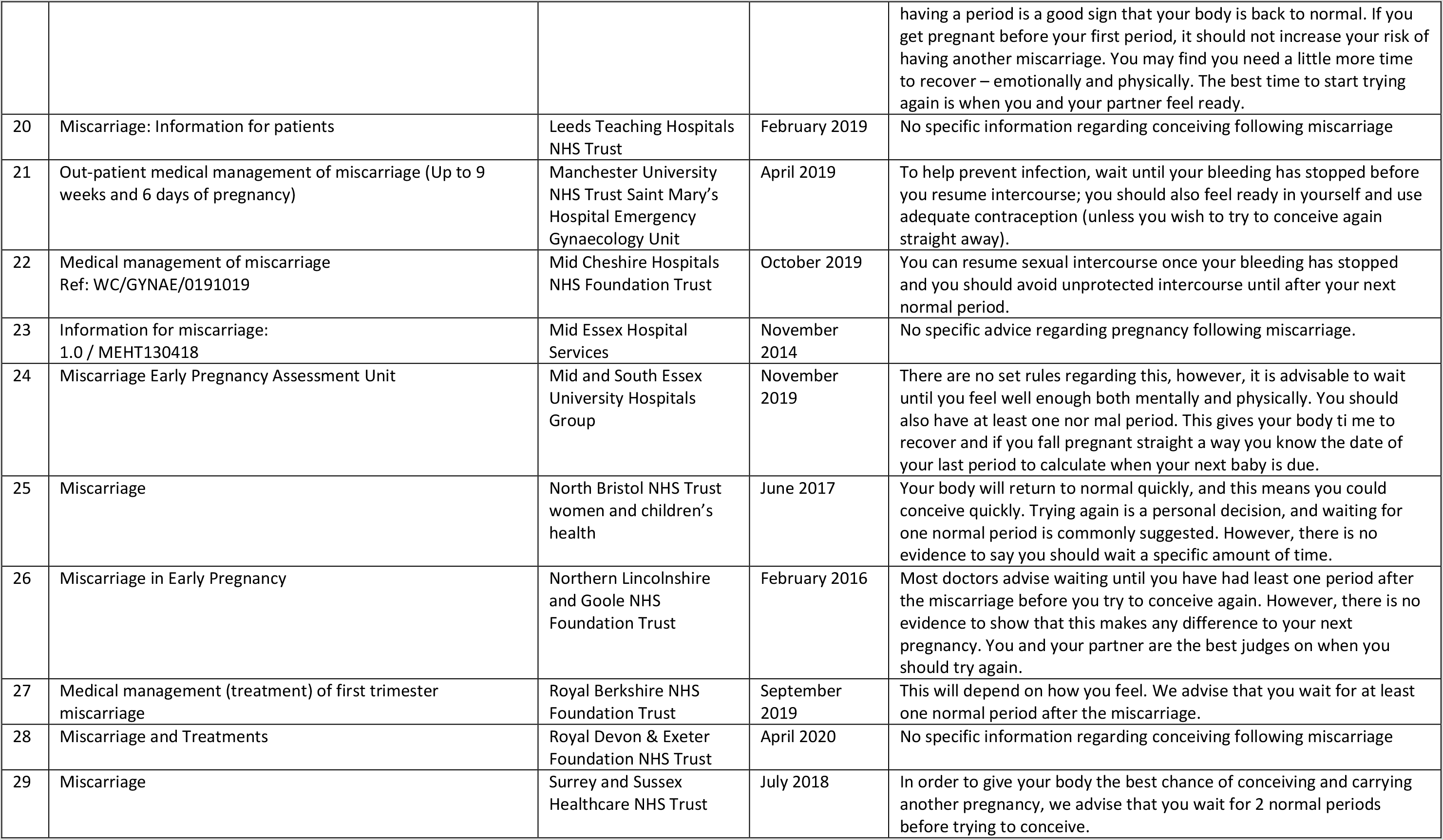
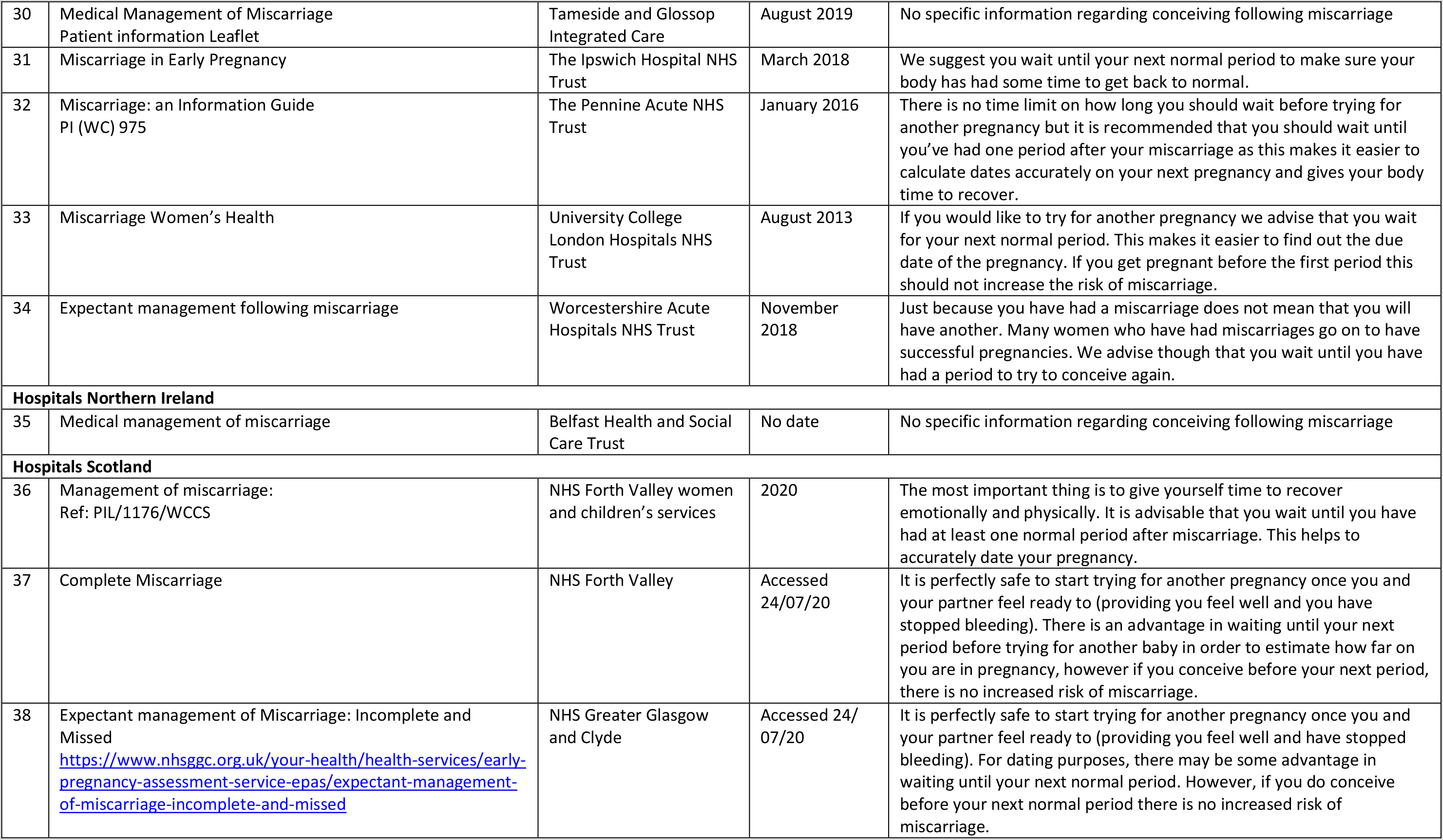
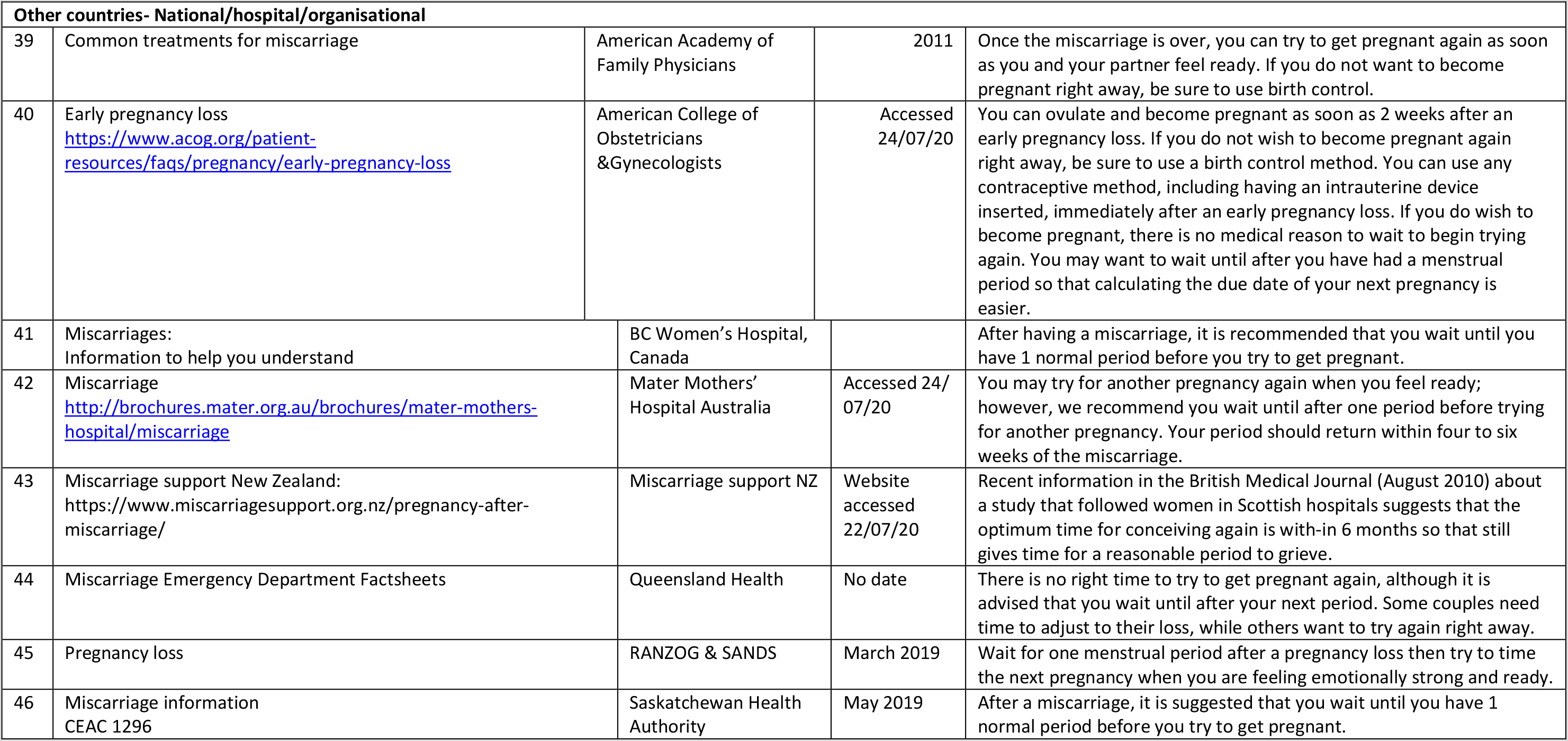
Characteristics of patient information leaflets regarding miscarriage issued by professional organisations and hospitals in the UK and other countries

## Discussion

### Principal findings

It was obvious from the large number of discussion threads found in relation to the timing of the next pregnancy following a miscarriage (n = 94) that this was an issue that women worry about. Analysis of online thread content in Mumsnet showed that women believed that there was no need to wait before attempting to conceive again following a miscarriage and that one should try to conceive again when feeling physically mentally and emotionally ready. Women posted about their frustrations at the conflicting advice they received and at the lack of professional sympathy. They felt that being told to wait before trying to conceive again is advice based on traditional guidelines. They also felt stressed by the pressures to conceive a successful pregnancy particularly by peer pressure and did not want to risk a further miscarriage by conceiving too early. In almost every thread analysed, women stated that they had been advised to wait for a normal period cycle before trying to conceive again and that the reason behind this was ease of calculating and dating the subsequent pregnancy, by the healthcare professional. In response to this, another post pointed out that, with advances in ultrasound scanning techniques, it was now possible to accurately date the next pregnancy even if it was conceived before a normal period. It was clear that the vast majority of women who had suffered a miscarriage and wanted to conceive again received an array of conflicting advice on when they can try again. From being told to wait for six months, to wait for one period cycle and also being told there is no requirement to wait at all, women have been challenged with having to make a decision regarding the optimum timing of the next pregnancy whilst coping with the grief and sense of loss. The implications of the received conflicting advice are that there is a loss of trust in the integrity of the medical profession and women appeared to put more faith in the advice from peers posting on the Mumsnet platform who have been through the same dilemma and had managed to have a successful pregnancy. (Carpenter et al., 2010) Numerous contributors in the thread posts have advised that utilising scans, investigations and other aids available, women should try and conceive after a miscarriage when they feel physically, emotionally and mentally ready to do so. The availability of internet access has made it possible for patients themselves to conduct their own research and spot any discrepancy in the advice given by health care professionals (NHS, 2019). This has had the effect of causing women to lose confidence in the medical profession; many have stated that they struggle to confide in their medical professional, as they doubt the advice they are being given is based on clear evidence. This is found to be a common phenomenon plaguing the medical profession and healthcare professionals have been urged to correct misinformation and resolve inconsistencies (BrainLine, 2019). Surveys have shown that the medical profession is the most trusted profession and patients trust the advice they are being given from medical professionals at face value. It is extremely important to ensure that this trust is not abused and the patient’s health and concerns should be of utmost importance (Daniels, 2019).

In contrast, our systematic web search for patient advice on timing of the next pregnancy following miscarriage revealed broadly consistent advice – to wait for a normal period before trying to conceive and/ or to try when the couples themselves felt ready. Despite the outdated guidance from WHO, none of the online advice suggested that couples wait for at least six months following a miscarriage before trying to conceive. In summary, the patient information leaflets identified from our web search all consistently advised couples that there was no clinical reason to delay a pregnancy following miscarriage. The consistent advice was in direct contrast to that reported by women themselves through the Mumsnet discussion platform. This contradiction may possibly be explained by the fact that all currently available patient information was dated after 2011, post publication of the article by Love et al (2010), indicating that the advice is updated regularly based on contemporaneous evidence.

It is important to acknowledge the chronological sequence of events in the context of this issue. The WHO guidelines published in 2005 was based on the best available evidence at that time. As is the problem with all guidelines there is a lag phase before new research evidence is incorporated into an updated guideline. Meanwhile the couples themselves appear have voted with their feet, opting to listen to their own bodies and empowered by peer advice and support to make the decision to conceive when they felt physically and emotionally ready. This decision appears to have been endorsed by current web-based support and information services rather than the other way around and in direct contradiction to WHO’s international guidelines.

### Findings in the context of the literature

Qualitative studies exploring the experiences and attitudes of women on pregnancy following a miscarriage have been carried out previously. Ockhuijsen et al, interviewed 24 women who had suffered a miscarriage and observed an overarching theme highlighting the need for ‘balancing between loss of control and searching for control’. The results of the study showed that most women reported feelings of anxiety and confusion following the miscarriage period and interval in conceiving again (Ockhuijsen et al., 2014). Bovin and Lancastle in their research on medical waiting periods, further displayed this theme of confusion and anxiety. Women experience anticipatory anxiety, as they are unsure of the outcomes in their subsequent pregnancies (Bolvin and Lancastle, 2010). Both these themes also figured strongly in our analysis of the Mumsnet discussion threads.

Andersson et al conducted a qualitative interview study on the feelings and emotions of pregnant women who have previously suffered a miscarriage. The research reported that women experienced an element of vulnerability and a lack of support from health care workers in the post miscarriage and subsequent conception phase. The women expressed a desire for sympathy and wish to have their questions answered adequately. Many of the women complained that they did not receive the support and understanding they expected in relation to their anxiety and concerns about being pregnant following a miscarriage. The study concluded that women believed that the presence of additional support during pregnancy aided in reducing the pre-delivery anxiety in those who have suffered from a miscarriage in the past (Andersson, Nilsson and Adolfsson, 2011). Strategies are required to help women in regaining control by advising women that help was available; and about scans, investigations, supplements and activities that are available to them during this difficult time (Ockhuijsen et al., 2014).

Qualitative studies reporting specifically about the timing of the next pregnancy following miscarriage are few and far between. In their questionnaire survey, Cuisiner et al report that conceiving again and the birth of a living child both reduced grief from a previous pregnancy loss (Cuisinier et al., 1996). An early conception following a miscarriage helped recovery and healing. They suggest that parents should no longer be advised to wait a specific time before conceiving again following a miscarriage and support them to make their own informed decision concerning the timing of the subsequent pregnancy taking account of their personal circumstances.

A number of factors affect the views and beliefs of women towards the interpregnancy interval after a miscarriage, such as fears from past experiences, the fear of trying too early following a miscarriage, stress and pressure from those around them and a lack of professional sympathy (Kinsey et al., 2013). According to Ellis et al, care following miscarriage is an area of healthcare, which is required to be revised and improved in order to ensure optimal outcomes and satisfaction for women who are trying to conceive (Ellis et al., 2016).

Gerber-Epstein et al (2011), in their interviews with Israeli women who have suffered a miscarriage in their first pregnancy, highlighted the influence of people and institutions with whom they were in contact during this period and formed an integral part of understanding the experience. These included the partner, the birth family and women who had had a similar experience. Encounters with friends who were pregnant, or attending social occasions produced feelings of distress, including envy and failure thus wishing to avoid such meetings. Similar experiences were reported by Bansen and Stevens (1992) - the taboo around miscarriage contributed to the inadequate support women received. Women somehow felt responsible for their miscarriages but healed better by accepting it unconditionally. They grieved their losses as they would mourn the deaths of other family members and all agreed that the experience was life-changing.

Other qualitative studies carried out with women following stillbirth or neonatal deaths often resonate with the findings from this research. Davis et al in their qualitative study based on individual interviews report that women were dissatisfied with advice from their doctors, saying that the timing of a subsequent pregnancy after a stillbirth was a personal decision. In their qualitative study using semi-structured interviews, Meaney et al (2016) reported about the fears of parents regarding the fear of another loss in a pregnancy following stillbirth and the expectations of additional hospital appointments and specialist care to prevent this recurrence. As with our own report, participants in the study by Lee et al (2013) described the importance of decisions regarding subsequent pregnancies being based on personal reasons, rather than thoughts of medical professionals or others.

### Strengths and limitations

This research study had many strengths, which include the dual nature of the study. The nature of the project was such that it allowed exploring the issue of the ideal interpregnancy interval following miscarriage from the perspectives of women themselves while comparing and contrasting this with the information that is currently given out by healthcare professionals. The searching of the web for patient information was robust and comprehensive, however it is possible that some information given out to women following a miscarriage may have been missed. Nevertheless, analysis from both a real-world as well as the stakeholders’ perspective was accomplished. (Carter, 2014).

Secondly, the anonymity of the discussion threads enabled women to express their views and beliefs in a non-threatening, supportive environment without any constraints or inhibitions. This is likely to allow organic discussion and views to be expressed, in comparison to an interview setting where participants may restrain their thoughts and feelings due to pressures and where the questions are designed in a specific format to achieve specific answers potentially biased by the interviewers’ own preconceptions (Open Textbooks for Hong Kong, 2019) Therefore this method of data collection from online threads allowed an unbiased perspective to be gleaned.

A further strength of the qualitative data analysis carried out is that it is of importance to the wider research community. As the research fills a gap in current knowledge surrounding the subject regarding the impact of policies and advice on couples, it is valuable to health researchers in terms of future policy development. This is highlighted by the fact that qualitative work is regarded as key in ‘people centered development’ and thus utilised in the development of recommendations and guidelines (Cornish, 2015).

A limitation of this study is that it may not be generalizable to a wider community. Mumsnet is said to be one of the most popular online forums in the UK for parents. Although Mumsnet states that it is ‘by parents, for parents’ (Mumsnet.com, 2019), it is heavily dominated by females, with an extremely low percentage of male users, men only make up 2-4% of core users on the forum (Mumsnet, 2009). Also, a survey conducted by Mumsnet found that users predominantly represent a higher socioeconomic status than population at large (Pedersen, 2016). Furthermore, as Mumsnet is an online-based forum it assumes that users have access to the Internet as well as digital literacy skills. Thus, this may automatically exclude large proportions of the population. Ultimately, these limitations may mean that the results of this study have to be restricted to the certain demographic population in question, and they may not be indicative of the views and beliefs of fathers, non-English speakers or non-users of the forum. Mumsnet does not publish demographic data regarding the gender of the respondents and pseudonyms are used by contributors. It was assumed that participants on the Mumsnet threads included here were mostly women who require advice and share their experiences regarding specific issues, based on the contents of their posts and their pseudonyms. However, it was challenging to determine whether respondents were mothers or fathers (Pedersen, 2015). Initially, the title of the research study was concerning the views and beliefs of couples, however this had to be revised given the fact that the Mumsnet data showed that the participants are majority female and it would be difficult to make a comment regarding ‘couples’’ thoughts.

Sample bias may have been in play in this study. It is likely that those that are strongly opinionated towards one position are more inclined to post on the discussion threads. Those with opposing views may be reluctant to post due to fear or criticism. Although, this is also viewed as a strength of the forum as those with strong views offer support and encouragement for other users who may have had similar experiences.

The systematic search of web-based information for patients was restricted to English language websites only and may therefore have missed differing advice given out in other languages. The search yielded predominantly UK based healthcare and charities with some information from the United States, Canada, Australia and New Zealand. None of the information was specifically tailored to women from low and middle-income countries where the use of web-based health information may be limited. We also could not access older patient information leaflets, which are taken down once updated information is made available. These could conceivably have contained the conflicting advice regarding optimum interpregnancy interval after miscarriage that the women posted about on Mumsnet.

### Recommendations for research and policy

The immense lack of direction, guidance and conflicting advice for women who suffered a miscarriage and are looking to conceive again warrants the need for further qualitative research to be conducted on this topic to enable formulation of contextually appropriate recommendations on the ideal interpregnancy interval following a miscarriage. Our research was mainly based on views expressed by women from developed countries with universal access to healthcare who were already empowered to carry out their own research. Online peer support facilitated decision making appropriate to personal circumstances even if this meant ignoring advice from healthcare professionals. As usual the voice of the minority who have limited healthcare access is not represented in this research; nor is there much evidence from low and middle-income countries regarding the optimum interpregnancy interval following miscarriage. Further research should address these knowledge gaps informing a clinically grounded international guideline universally acceptable to stakeholders and personal contexts.

## Conclusion

It was evident that women experience a sense of confusion and uncertainty about the ideal interpregnancy interval following a miscarriage. This may be due to conflicting advice from different healthcare professionals although this was not borne out by our search for web-based information on this topic. Women had found out that the advice to wait for a specific time period did not have any clinical basis and was based upon traditional norms and practices (Marinovich et al., 2019). Although women have complained about the conflicting messages received from healthcare personnel the evidence suggests that the current online information is consistent and based on updated evidence supporting individual decision-making. The analysis of the discussion threads portrayed that these women, who had often been victims of a lack of professional sympathy, had overwhelmingly concluded, contrary to the WHO guidelines, that there is no reason to wait following a miscarriage in trying to conceive again. Given the availability of investigations and scans, there is more personalised care available for the next pregnancy. Timing of the next pregnancy following miscarriage was a personal decision and to try to conceive again once the woman feels physically, mentally and emotionally ready to do so. A quote found during the analysis of the threads and chosen to be the title of the research study summarises the conclusion appropriately, ‘Your womb, Your choice’, displaying the belief that the ideal interpregnancy interval following a miscarriage is dependent on the wishes of the woman.

## Data Availability

The data used in this manuscript are freely available on the internet within www.mumsnet.comp

https://www.mumsnet.com/

## Notes

**Funding source:** This project formed part of FS’ MSc in Global Health and Management at University of Aberdeen; KL was supported by an institutional grant – REF Impact Support Award 18/19. The funders played no role in the collection, analysis or publication of the manuscript.

### Competing Interest Statement

The authors have declared no competing interest.

### Funding Statement

This project formed part of FS MSc in Global Health and Management at University of Aberdeen; KL was supported by an institutional grant REF Impact Support Award 18/19. The funders played no role in the collection, analysis or publication of the manuscript.

### Author Declarations

This research utilised publicly available online information and was therefore exempt from ethical approval.

